# Impact of the Russian invasion of Ukraine on the COVID-19 pandemic dynamics

**DOI:** 10.1101/2022.03.26.22272979

**Authors:** Igor Nesteruk

## Abstract

Full-scale invasion of Ukraine by Russian troops, which began on February 24, 2022, caused an unprecedented number of refugees, which in the ongoing COVID-19 pandemic could increase the number of patients. The recent pandemic dynamics in Ukraine, Poland, Germany and in the whole world was compared with the previous epidemic waves simulated with the use of the generalized SIR-model and corresponding parameter identification procedure. Since before the war the estimation of the number of infectious persons per capita in Ukraine 3.6 times exceeded the global figure, the increase of the number the new cases and the pandemic duration is expected. From the beginning of March 2022 the increase of the averaged number of new cases in Germany and worldwide is visible.

## Introduction

Russia’s full-scale criminal war in Ukraine has caused a real humanitarian catastrophe, the scale of which deserves appropriate assessment and punishment. As of March 23, 2022, more than 3.5 million Ukrainians have been forced to flee their homes and seek refuge abroad [1]. Such mass migration can lead to a significant increase in the number of COVID-19 cases.

In this paper we will try to reveal this trend by comparing the recent averaged numbers of new cases in Poland, Germany and the world with the relationships predicted for the previous epidemic waves with the use of generalized SIR-model [2], and a corresponding parameter identification procedure [3]. In particular, results of SIR simulations for 14^th^ epidemic wave in Ukraine [4, 5], 7^th^ global pandemic wave [4, 5], 4^th^ wave in Poland [6] and 5^th^ wave in Germany [6] will be used for comparison.

### Data, smoothing procedure and generalized SIR model

We will use the data set regarding the accumulated numbers of laboratory-confirmed COVID-19 cases *V*_*j*_ in Ukraine, Poland, Germany and the whole world from the COVID-19 Data Repository by the Center for Systems Science and Engineering (CSSE) at Johns Hopkins University (JHU), [7] (see Tables 1-4). It must be noted the JHU figures for Ukraine [7] are approximately 3% higher than the values reported by Ukrainian national sources [8, 9]. Due to the war the figures for Ukraine have not been updated after February 24, 2022 (see Table 1). The corresponding moments of time *t*_*j*_ (measured in days) are shown in Tables 1-4 for the period of November 2021 to March 2022. The JHU periodically updates its data sets for the previous moments of time. Here we will use the version of JHU file, corresponding to March 23, 2022. In particular, we will use the global data set presented in Table 2 which is slightly different from the previous version used in [4, 5].

**Table 1.** Cumulative numbers of laboratory-confirmed Covid-19 cases in Ukraine for the period of November 1, 2021 to February 24, 2022 according to JHU report on March 23, 2022, [7].

| Day in corresponding month of 2021 and 2022 | Number of cases in November 2021, $V_j$ | Number of cases in December 2021, $V_j$ | Number of cases in January 2022, $V_j$ | Number of cases in February 2022, $V_j$ |
| --- | --- | --- | --- | --- |
| 1 | 3073125 | 3619223 | 3852397 | 4287117 |
| 2 | 3093661 | 3633386 | 3854405 | 4323009 |
| 3 | 3118140 | 3647777 | 3856359 | 4363754 |
| 4 | 3146617 | 3661583 | 3858248 | 4408776 |
| 5 | 3174223 | 3668794 | 3862959 | 4452612 |
| 6 | 3200411 | 3673839 | 3869728 | 4481918 |
| 7 | 3218967 | 3683044 | 3877032 | 4506669 |
| 8 | 3233178 | 3692939 | 3880371 | 4542568 |
| 9 | 3253327 | 3705823 | 3883316 | 4582137 |
| 10 | 3277772 | 3717640 | 3885416 | 4625614 |
| 11 | 3303694 | 3728246 | 3890974 | 4668581 |
| 12 | 3328934 | 3733967 | 3898240 | 4708604 |
| 13 | 3353694 | 3738390 | 3908469 | 4735258 |
| 14 | 3369387 | 3746106 | 3919151 | 4753922 |
| 15 | 3381399 | 3754567 | 3929950 | 4785138 |
| 16 | 3398913 | 3764485 | 3936582 | 4818112 |
| 17 | 3418792 | 3773700 | 3941923 | 4853339 |
| 18 | 3440602 | 3781506 | 3950774 | 4890332 |
| 19 | 3461873 | 3785395 | 3963917 | 4923680 |
| 20 | 3481347 | 3788209 | 3982738 | 4943428 |
| 21 | 3493203 | 3794490 | 4003280 | 4959461 |
| 22 | 3501815 | 3801079 | 4026198 | 4986161 |
| 23 | 3515641 | 3808612 | 4042152 | 5012980 |
| 24 | 3530969 | 3815440 | 4055643 | 5040518 |
| 25 | 3548842 | 3820891 | 4075351 | No data |
| 26 | 3565644 | 3823879 | 4100292 | No data |
| 27 | 3580671 | 3825917 | 4133396 | No data |
| 28 | 3588916 | 3828336 | 4168560 | No data |
| 29 | 3595410 | 3833952 | 4206731 | No data |
| 30 | 3606622 | 3840041 | 4232143 | No data |
| 31 | - | 3847226 | 4255206 | No data |

**Table 2.** Cumulative numbers of laboratory-confirmed Covid-19 cases in the world for the period of November 1, 2021 to March 22, 2022 according to JHU report on March 23, 2022, [7].

| Day in<br>corres-<br>ponding<br>month of<br>2021 and<br>2022 | Number of<br>cases in<br>November<br>2021,<br>$V_j$ | Number of<br>cases in<br>December<br>2021,<br>$V_j$ | Number of<br>cases<br>in January<br>2022,<br>$V_j$ | Number of<br>cases<br>in February<br>2022,<br>$V_j$ | Number of<br>cases<br>in March<br>2022,<br>$V_j$ |
| --- | --- | --- | --- | --- | --- |
| 1 | 247861854 | 263889719 | 289933905 | 382299452 | 438729232 |
| 2 | 248285819 | 264595861 | 290853823 | 385517611 | 440402821 |
| 3 | 248824382 | 265313142 | 293243463 | 388702973 | 442305215 |
| 4 | 249347990 | 265827599 | 295774478 | 391613541 | 444024074 |
| 5 | 249868870 | 266267879 | 298356951 | 393822977 | 445417573 |
| 6 | 250296481 | 266856872 | 301009468 | 395644279 | 446578954 |
| 7 | 250646490 | 267554327 | 303949573 | 398195810 | 448089716 |
| 8 | 251125070 | 268225326 | 306104961 | 401031723 | 449944357 |
| 9 | 251611936 | 268955015 | 308126442 | 403489136 | 451829923 |
| 10 | 252200453 | 269640522 | 311304268 | 406257339 | 453652969 |
| 11 | 252720115 | 270140464 | 314409815 | 408645179 | 455486874 |
| 12 | 253314581 | 270580682 | 317868825 | 410471795 | 457144094 |
| 13 | 253745142 | 271204631 | 321059669 | 411926999 | 458350633 |
| 14 | 254109339 | 271864323 | 324360279 | 413724219 | 460085135 |
| 15 | 254646560 | 272609345 | 326824415 | 415507791 | 461950527 |
| 16 | 255173747 | 273346817 | 328983101 | 417927182 | 464203918 |
| 17 | 255808066 | 274079144 | 331690684 | 419963810 | 466258201 |
| 18 | 256423122 | 274662171 | 335494877 | 421905788 | 468133052 |
| 19 | 257043551 | 275144793 | 339610692 | 423458977 | 469713452 |
| 20 | 257525406 | 275886668 | 343323721 | 424728142 | 470737068 |
| 21 | 257929555 | 276684579 | 347152734 | 426187929 | 472116383 |
| 22 | 258547892 | 277593721 | 349889102 | 427995572 | 474114550 |
| 23 | 259161029 | 278589936 | 352316289 | 429938984 |  |
| 24 | 259834033 | 279450592 | 355809402 | 431697291 |  |
| 25 | 260431579 | 280147825 | 359513666 | 433290944 |  |
| 26 | 261030829 | 280727880 | 363240730 | 434650622 |  |
| 27 | 261492296 | 282000616 | 366954227 | 435736709 |  |
| 28 | 261901725 | 283336850 | 370568546 | 437165670 |  |
| 29 | 262553270 | 285063022 | 373234356 | - |  |
| 30 | 263176553 | 287004720 | 375503393 | - |  |
| 31 | - | 288731494 | 379089774 | - |  |

**Table 3.** Cumulative numbers of laboratory-confirmed Covid-19 cases in Poland for the period of November 1, 2021 to March 22, 2022 according to JHU report on March 23, 2022, [7].

| Day in<br>corres-<br>ponding<br>month<br>of 2021<br>and<br>2022 | Number of<br>cases in<br>November<br>2021,<br>$V_j$ | Number of<br>cases in<br>December<br>2021,<br>$V_j$ | Number of<br>cases in<br>January<br>2022,<br>$V_j$ | Number of<br>cases in<br>February<br>2022,<br>$V_j$ | Number of<br>cases in<br>March<br>2022,<br>$V_j$ |
| --- | --- | --- | --- | --- | --- |
| 1 | 3030151 | 3569137 | 4120248 | 4925270 | 5680034 |
| 2 | 3034668 | 3596491 | 4127428 | 4981321 | 5694767 |
| 3 | 3045102 | 3623452 | 4133851 | 5035796 | 5708827 |
| 4 | 3060613 | 3649027 | 4145518 | 5083332 | 5721316 |
| 5 | 3076518 | 3671421 | 4162715 | 5129080 | 5734041 |
| 6 | 3091713 | 3684671 | 4179292 | 5163780 | 5741739 |
| 7 | 3104220 | 3704040 | 4191193 | 5188184 | 5747322 |
| 8 | 3111534 | 3732589 | 4202090 | 5223507 | 5760498 |
| 9 | 3125179 | 3760048 | 4213197 | 5270363 | 5774938 |
| 10 | 3143725 | 3785036 | 4220984 | 5312450 | 5788363 |
| 11 | 3162804 | 3808798 | 4232386 | 5348224 | 5799996 |
| 12 | 3175769 | 3828248 | 4248559 | 5379551 | 5811109 |
| 13 | 3190067 | 3839625 | 4265433 | 5401615 | 5818687 |
| 14 | 3204515 | 3857085 | 4281482 | 5415088 | 5823982 |
| 15 | 3214023 | 3881349 | 4298375 | 5437343 | 5836672 |
| 16 | 3230634 | 3903445 | 4313036 | 5466198 | 5851147 |
| 17 | 3254875 | 3923472 | 4323482 | 5495432 | 5863414 |
| 18 | 3279787 | 3942864 | 4343130 | 5519411 | 5875072 |
| 19 | 3303046 | 3958840 | 4373718 | 5540302 | 5885446 |
| 20 | 3326464 | 3968450 | 4406553 | 5553989 | 5891140 |
| 21 | 3345388 | 3982257 | 4443217 | 5563446 | 5895304 |
| 22 | 3357763 | 4000270 | 4484095 | 5582217 | 5905463 |
| 23 | 3377698 | 4017420 | 4518218 | 5602680 |  |
| 24 | 3406129 | 4032796 | 4547315 | 5620946 |  |
| 25 | 3434272 | 4043585 | 4584360 | 5637646 |  |
| 26 | 3461066 | 4049838 | 4637776 | 5651596 |  |
| 27 | 3487254 | 4054865 | 4695435 | 5660493 |  |
| 28 | 3507828 | 4064715 | 4752700 | 5667054 |  |
| 29 | 3520961 | 4080282 | 4804390 | - |  |
| 30 | 3540061 | 4094608 | 4852677 | - |  |
| 31 | - | 4108215 | 4886154 | - |  |

**Table 4.**
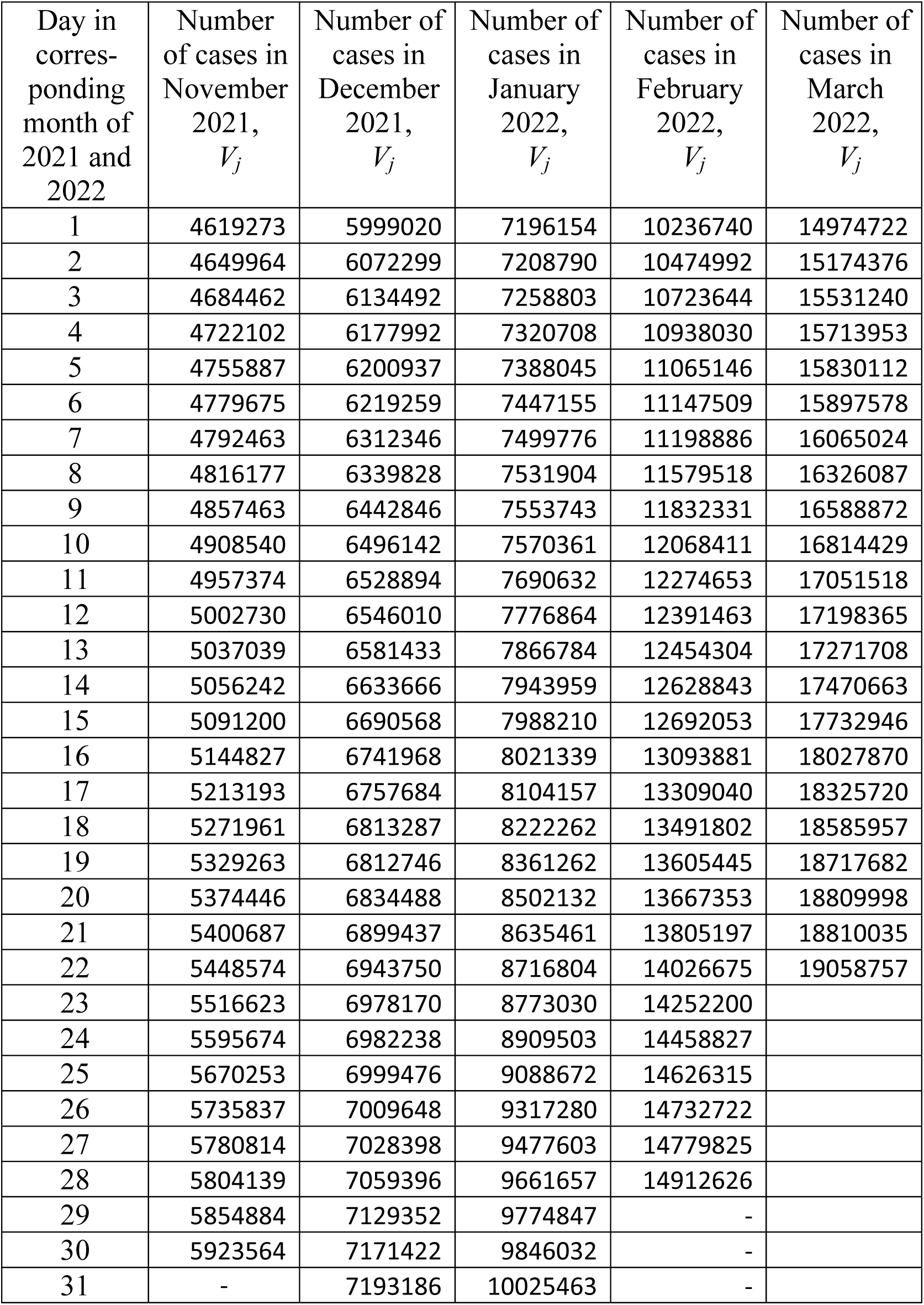
Cumulative numbers of laboratory-confirmed Covid-19 cases in Germany for the period of November 1, 2021 to March 22, 2022 according to JHU report on March 23, 2022, [7].

The generalized SIR-model relates the numbers of susceptible *S(t)*, infectious *I(t)* and removed persons *R(t)* versus time *t* for a particular epidemic wave *i*, [2]. The exact solution of the set of non-linear differential equations uses the function *V* (*t*) = *I* (*t*) + *R*(*t*), corresponding to the number of victims or the cumulative laboratory-confirmed number of cases [2]. Its derivative *dV/dt* yields the estimation of the average daily number of new cases. When the registered number of victims *V*_*j*_ is a random realization of its theoretical dependence, the exact solution presented in [2] depends on five parameters. The details of the optimization procedure for their identification can be found in [3].

Since daily numbers of new cases are random, we will use the smoothed number of accumulated cases and its numerical derivative to estimate the smoothed number of new daily cases (see details in [10]).

## Results and discussion

The optimal values of the general SIR model and other characteristics of the 14th epidemic wave in Ukraine [4, 5], the 4^th^ wave in Poland [6], the 5^th^ wave in Germany [6] and the 7^th^ wave in the whole world [4, 5] are listed in Table 5. Corresponding SIR curves are shown in Figs. 1 and 2. The laboratory confirmed accumulated numbers of COVID-19 cases *V*_*j*_ (Tables 1-4) are shown by “circles” (data used for SIR simulations) and “stars” (data used for control the results of calculations). “Crosses” represent the averaged daily numbers of new COVID-19 cases calculated with the use of the *V*_*j*_ values.

**Table 5.** Optimal values of parameters and other characteristics of the COVID-19 pandemic waves in Ukraine, Poland, Germany and the world.

| Characteristics | 14<sup>th</sup> epidemic wave<br>in Ukraine,<br>$i=14$,<br>[4, 5] | 4<sup>th</sup> epidemic wave<br>in Poland,<br>$i=4$,<br>[6] | 5<sup>th</sup> epidemic wave<br>in Germany,<br>$i=5$,<br>[6] | 7<sup>th</sup> epidemic wave<br>in the whole world,<br>$i=7$,<br>[4, 5] |
| --- | --- | --- | --- | --- |
| Time period<br>taken for<br>calculations, $T_{ci}$ | January 22 –<br>February 4, 2022 | November 22 –<br>December 5, 2021 | November 22 –<br>December 5, 2021 | January 22 –<br>February 4, 2022 |
| $I_i$ | 64,676.7037685832 | 182,880.050730977 | 175,036.042911984 | 11,190,879.9375884 |
| $R_i$ | 3,956,648.86765999 | 3,181,514.23498331 | 5,301,466.81423087 | 337,916,505.348126 |
| $N_i$ | 5,678,291.86675200 | 5,410,976 | 10,300,000 | 815,388,678.72 |
| $\nu_i$ | 1,064,719.47680477 | 1,574,583.94143908 | 4,370,737.14176735 | 453,226,448.070179 |
| $\alpha_i$ | 1.7392861614679e-7 | 5.96394337877141e-08 | 6.8098348012594e-08 | 6.51375439773073e-10 |
| $\rho_i$ | 0.185185185185185 | 0.0939072947186539 | 0.297639978951643 | 0.295220576928501 |
| $1/\rho_i$ | 5.4 | 10.6488 | 3.35976371024561 | 3.38729776360470 |
| $r_i$ | 0.998361163644707 | 0.998395644300319 | 0.996581480894305 | 0.999439451877399 |
| $S_{i\infty}$ | 552,746 | 854,850 | 3,186,419 | 359,119,917 |
| $V_{i\infty}$ | 5,125,546 | 4,556,126 | 7,113,581 | 456,268,762 |
| Final day of the<br>epidemic wave | December 2023 | September 2025 | April 2023 | April 2085 |

**Fig. 1.**
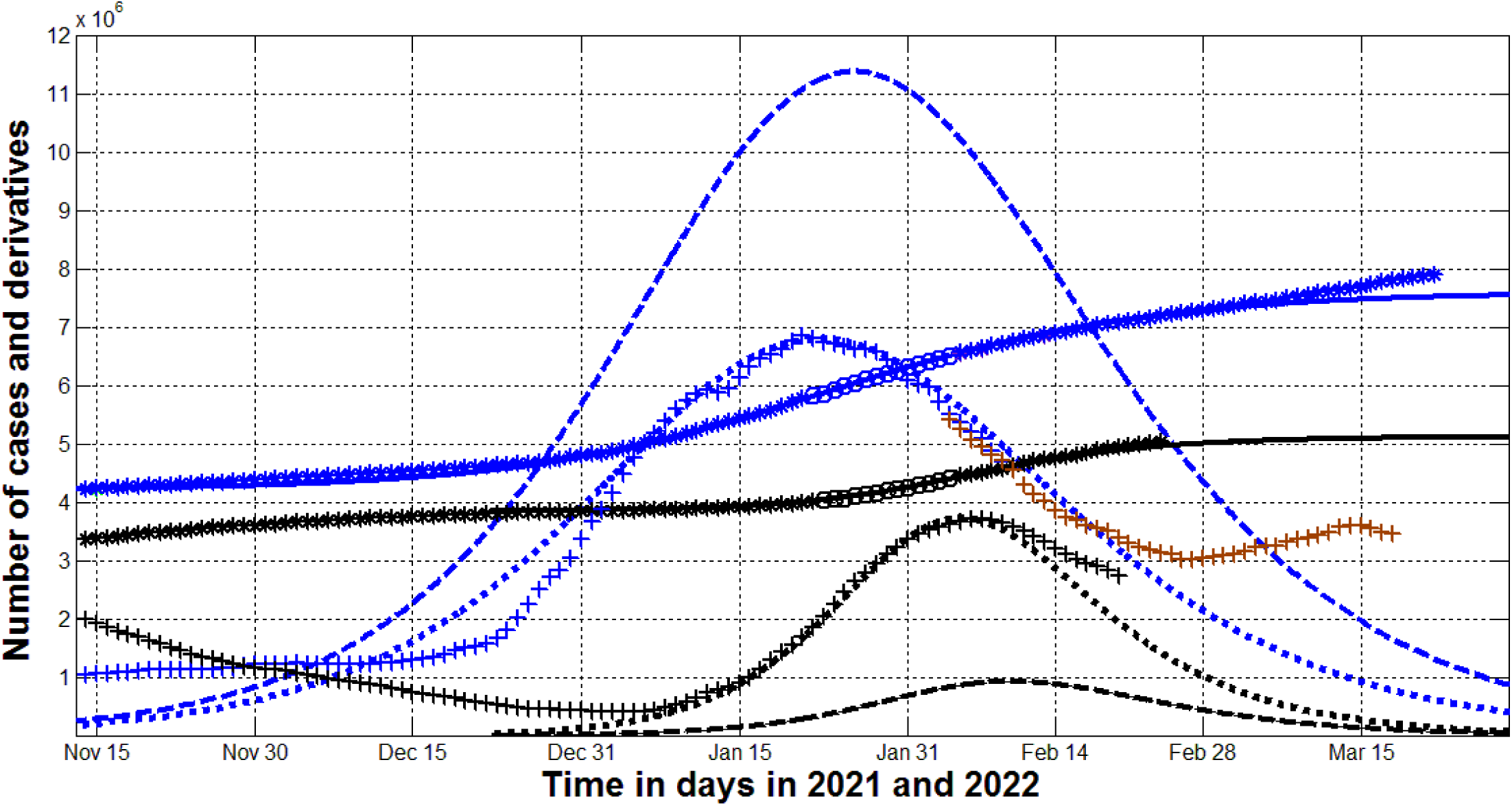
The omicron waves and further pandemic dynamics in Ukraine and in the whole world. The results of SIR simulations of the 14th wave in Ukraine are shown by black lines. Blue lines represent the 7^th^ pandemic wave in the world. Numbers of victims *V(t)=I(t)+R(t)* – solid lines (for the world divided by 60); numbers of infected and spreading *I(t)* (multiplied by 5 for Ukraine) – dashed; derivatives *dV/dt*, multiplied by 100 for Ukraine and by 2 for the world) – dotted. “Circles” correspond to the accumulated numbers of cases registered during the periods of time taken for SIR simulations (for the world divided by 60). “Stars” corresponds to *V*_*j*_ values beyond these time periods (for the world divided by 60). “Crosses” show the numerical first derivative multiplied by 100 for Ukraine and by 2 for the world. Black markers correspond to Ukraine ([7], Table 1), blue and brown “crosses” - for the world (different versions of JHU datasets [7], Table 2).

**Fig. 2.**
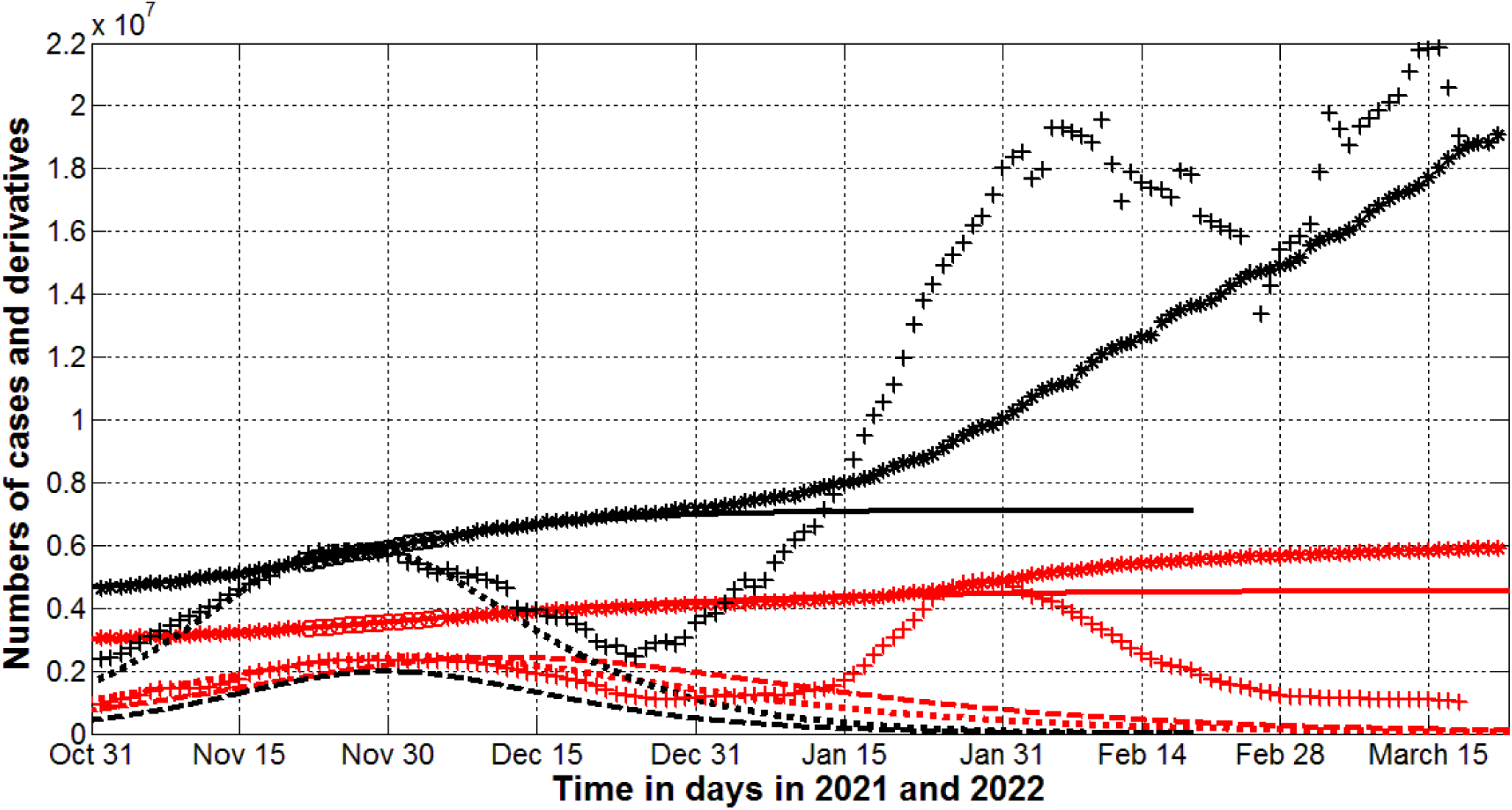
The COVID-19 pandemic waves in Poland (red) and Germany (black) in 2021 and 2022. The results of SIR simulations of the 4th wave in Poland and the 5th wave in Germany are shown by red and black lines, respectively. Numbers of victims *V(t)=I(t)+R(t)* – solid lines; numbers of infected and spreading *I(t)* multiplied by 10 – dashed; derivatives *dV/dt* (multiplied by 100) – dotted. “Circles” correspond to the accumulated numbers of cases registered during the period of time taken for SIR simulations. “Stars” correspond to *V*_*j*_ values beyond this time period. “Crosses” show the averaged daily number of new cases (calculated with the use of datasets presented in Tables 3, 4 and multiplied by 100).

“Stars” and “crosses” in Fig. 1 illustrate that before the war the accumulated number of cases (“stars”) and the averaged daily numbers of new cases (“crosses”) followed the corresponding theoretical solid and dotted lines. In March 2022 the real global dynamics started to deviate form the theoretical blue solid and dotted curves. In particular, the saturation level of the 7^th^ pandemic wave *V*_*i*∞_ =456,268,762 (see the last column of Table 5) is already exceeded. The increase in the global daily numbers of new cases (see brown “crosses”) can be explained by the mass migration from Ukraine. As of March 23, 2022, more than 3.5 million Ukrainians were forced to flee abroad [1].

To estimate the possible impact of this humanitarian disaster, let us calculate the probability of meeting an infectious person in Ukraine with the use of simple formula, [10]:

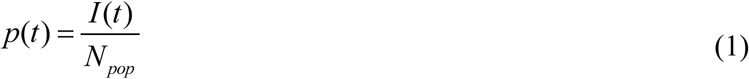

where *N*_*pop*_ is the volume of population. As of February 24, 2022 the numbers of people spreading the infection *I(t)* were around 100, 000 in Ukraine and 5 million in the whole world (see dashed lines in Fig. 1). Since before the war, the population of Ukraine was 178 times less than global figure, the probability of meeting an infected person in Ukraine was 3.6 times higher (according to eq. (1)). It means that forced mass emigration of Ukrainians could cause an increase in the number of new cases in the world. Brown crosses in Fig. 1 illustrate this fact. It is worth noting that after March 15, 2022 the growth stopped, which can be explained by a decrease in the flow of refugees.

Let us consider the situation in the Poland, which has accepted more than 2 million Ukrainian refugees [1]. In March 2022, the decline in the number of new cases slowed down (see red “crosses” in Fig. 2). The lack of an increase in the number of new cases in Poland can be explained by the approximately same probability of meeting an infected person with Ukraine.

Unfortunately, we have results of SIR simulations only for the 4^th^ wave in Poland (shown by red lines in Fig. 2, [6]). In January 2022 a new Omicron wave started in this country and the daily numbers of new cases (red “crosses” in Fig.2) became much higher than the theoretical estimation for the previous wave (the red dotted line). The maximum values of *I(t)* were approximately 200,000 both for Ukraine and Poland (see the black dashed line in Fig.1 and the red dashed line in Fig.2). Since the populations of these countries are also close, we can expect the close values for the probabilities of meeting of an infectious person (according to eq. (1)). Thus, the huge number of Ukrainian refuges did not change much the epidemic dynamics in Poland.

In early 2022, when a new powerful epidemic wave began in Germany, the number of infected in this country was about 4 times less than in Poland (compare black and red dashed lines in Fig. 2). Taking into account the difference in population size, one can expect about eight times less chance of meeting an infected person in Germany. Therefore, refugees from Ukraine could significantly increase the number of new cases in Germany in March 2022. Black “crosses” in Fig. 2 illustrate this fact.

## Conclusions

The huge number of Ukrainian refugees has increased the global number of new COVID-19 cases. Deterioration of epidemic dynamics should be expected in countries with a small number of infected per capita.

## Data Availability

All data produced in the present work are contained in the manuscript

## Acknowledgement

The author is grateful to Oleksii Rodionov for his help in collecting and processing data.

